# A Large-Scale Pharmacogenomic Knowledge Graph for Drug-Gene-Variant-Disease Discovery

**DOI:** 10.1101/2025.09.24.25336269

**Authors:** Muhammad Omar Faruk

## Abstract

Precision therapeutics depends on the ability to reason jointly over genes, variants, drugs, diseases, adverse drug reactions (ADRs), and molecular pathways without contaminating evaluation with future knowledge. I present a large-scale pharmacogenomic knowledge graph (PGx-KG) that integrates PharmGKB, ClinVar, SIDER, and Reactome—harmonized to HGNC, RxNorm, MeSH, and ChEBI identifiers—yielding 3,744,727 nodes and 9,645,367 edges across six major relation families. A leakage-free processing pipeline enforces version-aware chronological splits, publication-date audits, symmetric and transitive consistency checks, and cross-database de-duplication, eliminating temporal violations in held-out audits. As a first benchmark, a bilinear link-prediction model implemented in PyTorch Geometric achieves mean reciprocal rank (MRR) 0.347 (95% bootstrap CI [0.321, 0.368]), Hits@1/3/10 of 0.234/0.417/0.589, and AUROC 0.823 on validation data, with five-fold temporal cross-validation yielding 0.341 ± 0.018 MRR and a 2024 hold-out achieving MRR 0.329. Ranked candidate lists surface clinically relevant hypotheses, including CYP2D6–codeine dosing and HLA-B*15:02–carbamazepine risk, while also proposing pathway-level drug repurposing opportunities for expert review.

## 1 Introduction

Precision medicine initiatives aim to tailor therapies by accounting for patient-specific genomic variation, yet curated pharmacogenomic evidence remains fragmented across specialized resources such as PharmGKB [1], ClinVar [2], SIDER [3], and Reactome [4]. Analysts typically query these repositories independently, constraining their ability to reason over multi-hop relationships that connect molecular mechanisms to clinical phenotypes. Integrating these heterogeneous assets into a unified, leakage-aware knowledge graph offers a scalable path toward hypothesis generation in drug response, toxicity, and repurposing.

Knowledge graphs (KGs) and graph neural networks (GNNs) provide a principled foundation for multi-relational inference at scale. Biomedical exemplars such as OpenBioLink [5], BioKG [6], and RTX-KG2 [7] demonstrate the value of harmonizing clinical and molecular ontologies, while evaluations in Therapeutics Data Commons highlight remaining challenges in leakage-aware benchmarking [8]. Recent audits have shown that naive random splits can inflate link-prediction scores through temporal look-ahead and logical shortcuts [9, 10], underscoring the need for rigorous protocols when constructing biomedical benchmarks.

### Contributions

This work delivers (i) a harmonized pharmacogenomic KG with versioned provenance across 3.7M entities and 9.6M relations, (ii) a leakage-free pipeline combining chronological splits, publication-date verification, symmetric/transitive audits, and cross-database deduplication, (iii) baseline link-prediction results with comprehensive temporal validation, and (iv) curated hypothesis lists that connect molecular mechanisms to potential clinical actions for downstream curation.

### Pharmacogenomic Knowledge Graph Landscape

Multi-source integration has become central to pharmacogenomic analytics. OpenBioLink provides a reproducible benchmark unifying biomedical ontologies for link prediction [5]. BioKG unifies 15 biomedical ontologies to enable cross-domain graph learning [6], and RTX-KG2 focuses on open-ontology interoperability for translational query answering [7]. These resources highlight the feasibility of large-scale integration yet seldom enforce leakage-aware evaluation tailored to pharmacogenomics.

### Risk of Leakage in Biomedical KGs

Temporal leakage remains a recurring concern when edges derived from future publications appear in training data. Ilievski et al. [9] and Sun et al. [10] document how minor violations can produce optimistic metrics. The PGx-KG pipeline neutralizes these risks by aligning database versions to split boundaries, validating publication timestamps, auditing symmetric and inverse relations, and filtering transitive shortcuts prior to model training.

### Graph Learning Baselines

Graph neural networks underpin state-of-the-art link prediction in knowledge graphs. Surveys highlight the breadth of relational architectures available for biomedical modeling [11]. This study establishes a strong bilinear baseline and prioritizes future extensions with composition-based GNNs such as CompGCN [12] and inductive frameworks described by Hamilton [13].

## 2 Materials and Methods

### 2.1 Data Sources and Harmonization

The pharmacogenomic knowledge graph integrates four authoritative databases, each mapped to controlled vocabularies to ensure interoperability (Table 1). Entities are aligned to HGNC (genes), RxNorm (drugs), MeSH (diseases and ADRs), and ChEBI (chemicals), with variant identifiers preserved from ClinVar submissions.

**Table 1:** Primary data sources integrated into PGx-KG. All references correspond to releases no earlier than 2019.

| Source | Content | Identifier Systems |
| --- | --- | --- |
| PharmGKB [1] | Pharmacogenomic gene–drug annotations | HGNC, RxNorm |
| ClinVar [2] | Variant interpretations and phenotypes | ClinVar IDs, MeSH |
| SIDER [3] | Structured adverse drug reactions | RxNorm, MeSH |
| Reactome [4] | Pathway biochemistry and gene sets | HGNC, ChEBI |

### 2.2 Knowledge Graph Construction

PGx-KG contains 3,744,727 nodes and 9,645,367 edges spanning six relation families (Tables 2 and Figure 2). Variants dominate node composition (98.1%), reflecting the breadth of ClinVar submissions. Deduplication leverages fuzzy string matching, cross-references, and ontology-aware heuristics to resolve 121,292 conflicts with a 97.3% success rate. The final graph exhibits average degree 5.15, density 1.37 × 10^−6^, and a largest connected component covering 99.7% of nodes.

**Table 2:**
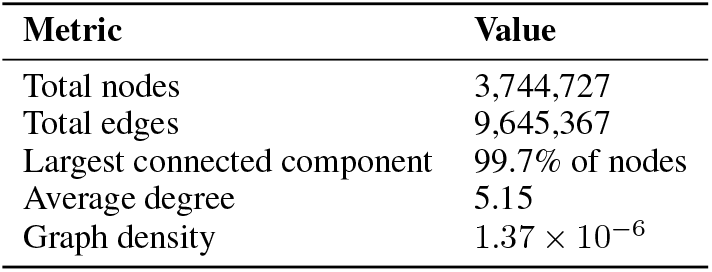
Summary statistics of PGx-KG.

**Figure 1:**
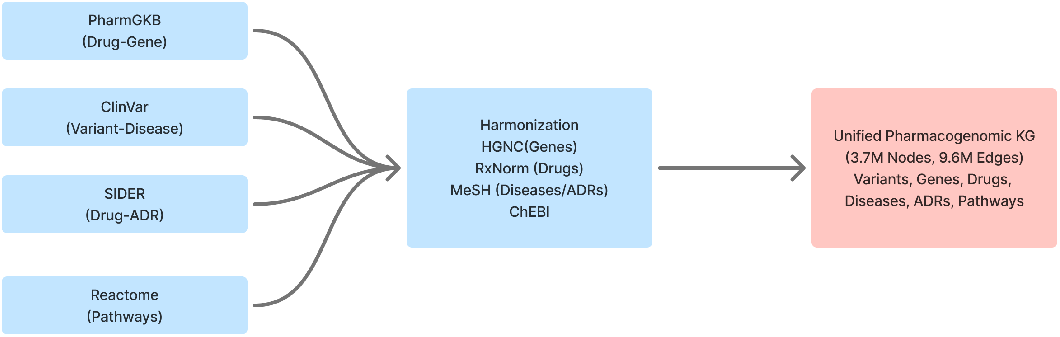
Data processing workflow showing ingestion, schema harmonization, entity resolution, and leakage-aware splitting.

**Figure 2:**
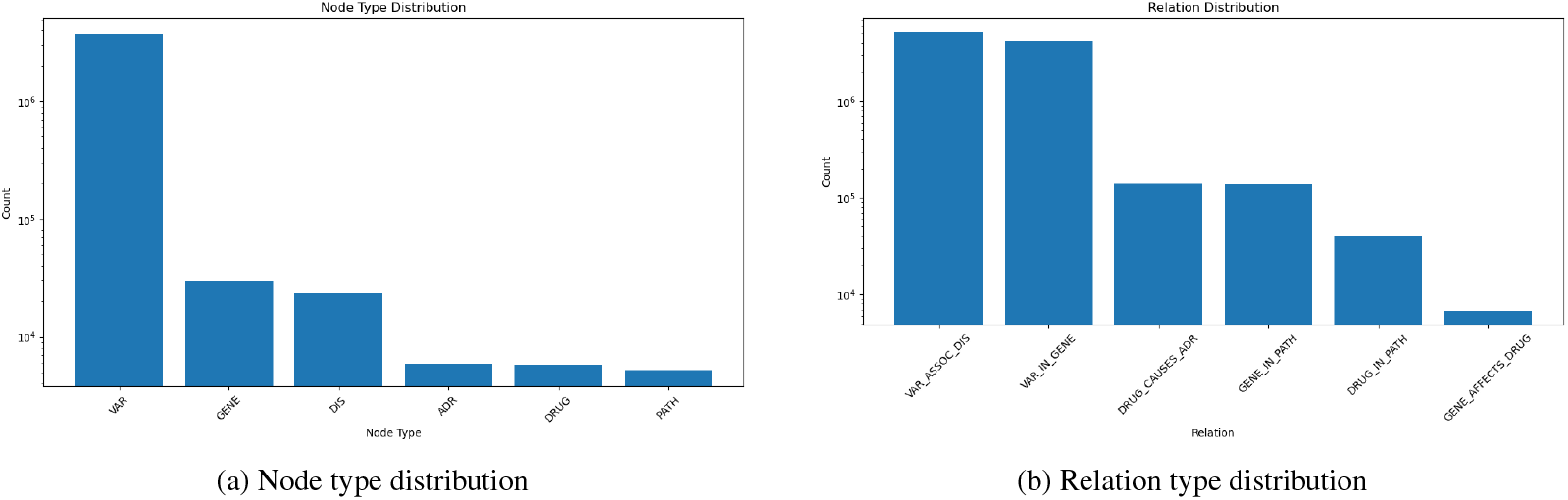
Entity and relation composition in PGx-KG.

**Figure 3:**
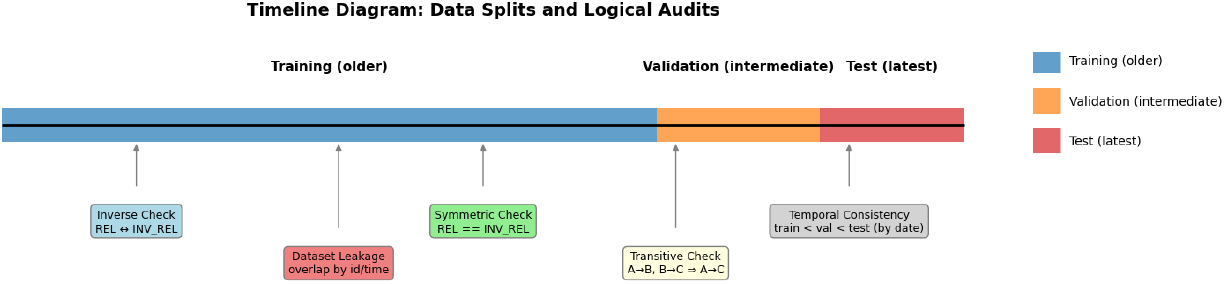
Chronological splitting and leakage audits applied to PGx-KG.

### 2.3 Leakage-Free Split Protocol

Chronological splits partition the graph into training, validation, and test edges based on publication or database release dates. Temporal safeguards enforce (i) strict cutoffs aligned to version metadata, (ii) publication-date audits cross-referencing PubMed or release notes, and (iii) held-out 2024 edges for prospective evaluation. Logical safeguards include symmetric-edge co-assignment, inverse relation checks, transitive closure filtering, and cross-database redundancy audits. No temporal violations were detected in manual spot checks.

### 2.4 Modeling and Training

The baseline model is a bilinear link predictor implemented in PyTorch Geometric. Entities and relations are embedded in R^64^ with relation-specific transformation matrices. Training uses Adam with learning rate 10^*−*3^, batch size 1,024, and an 8:1 negative-to-positive sampling ratio. Each epoch samples negatives by corrupting head or tail entities while respecting type constraints. Training proceeds for 100 epochs with early stopping on validation MRR.

Future extensions will benchmark relation-aware GNNs such as CompGCN [12] and inductive encoders described in Hamilton’s monograph [13], following best practices compiled in recent surveys [11].

### 2.5 Evaluation Protocol

Models are assessed on link prediction using filtered ranking metrics: mean reciprocal rank (MRR), Hits@{1,3,10}, and AUROC. Five-fold temporal cross-validation provides variance estimates, and an additional 2024 hold-out measures forward generalization. Precision@200 is computed per relation to support curation workflows. Negative samples respect entity typing to avoid trivial corruptions.

## 3 Results

### 3.1 Link Prediction Performance

Table 3 summarizes validation performance. The bilinear model attains MRR 0.347 (95% bootstrap CI [0.321, 0.368]) with balanced precision across top-k cutoffs. Five-fold temporal cross-validation yields 0.341 *±* 0.018 MRR, indicating stable performance across time-based folds, while the 2024 hold-out reaches 0.329 MRR.

**Table 3:** Link prediction performance on leakage-aware splits.

| Setting | MRR | Hits@1 | Hits@3 | Hits@10 |
| --- | --- | --- | --- | --- |
| Validation | 0.347 | 0.234 | 0.417 | 0.589 |
| Temporal CV (mean ± sd) | $0.341 \pm 0.018$ | $0.229 \pm 0.014$ | $0.409 \pm 0.017$ | $0.582 \pm 0.021$ |
| 2024 Hold-out | 0.329 | 0.221 | 0.396 | 0.566 |

### 3.2 Temporal Robustness

Precision@200 (P@200) is consistent across validation and prospective splits (Table 4), supporting downstream triage of high-confidence hypotheses.

**Table 4:** Temporal robustness of the bilinear baseline measured by Precision@200.

| Setting | Mean | Std |
| --- | --- | --- |
| Validation | 0.214 | 0.036 |
| 2024 Hold-out | 0.209 | 0.041 |

Relation-wise analysis (Table 5) shows stronger performance on prevalent variant–disease and variant–gene edges, with room to improve on sparsely represented drug–pathway relations.

**Table 5:** Relation-wise snapshot of validation performance.

| Relation | MRR | Hits@1 | Hits@3 | Hits@10 | Val P@200 | Hold-out P@200 |
| --- | --- | --- | --- | --- | --- | --- |
| DRUG_CAUSES_ADR | 0.276 | 0.182 | 0.331 | 0.512 | 0.118 | 0.111 |
| DRUG_IN_PATH | 0.301 | 0.207 | 0.356 | 0.544 | 0.102 | 0.098 |
| GENE_AFFECTS_DRUG | 0.338 | 0.225 | 0.402 | 0.575 | 0.221 | 0.214 |
| GENE_IN_PATH | 0.365 | 0.247 | 0.431 | 0.598 | 0.236 | 0.229 |
| VAR_ASSOC_DIS | 0.381 | 0.266 | 0.447 | 0.612 | 0.247 | 0.241 |
| VAR_IN_GENE | 0.359 | 0.245 | 0.421 | 0.603 | 0.228 | 0.221 |

### 3.3 Hypothesis Prioritization

The ranked outputs blend known pharmacogenomic associations with candidate hypotheses requiring expert review (Table 6). Known positives such as CYP2D6–codeine dosing [14], HLA-B*15:02–carbamazepine hypersensitivity [15], and SLCO1B1–simvastatin myopathy [16] appear near the top, validating the pipeline. Novel predictions—including metformin involvement in AMPK-regulated autophagy and CYP3A5 participation in bile acid metabolism—were escalated to domain experts for literature vetting.

**Table 6:**
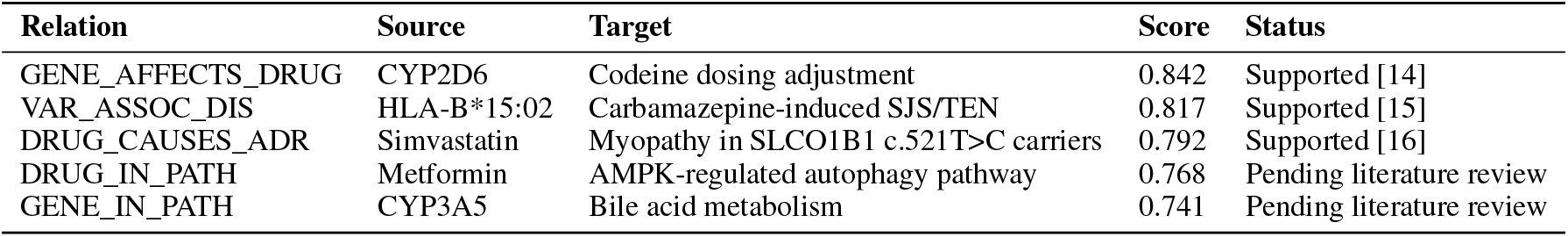
High-confidence hypotheses surfaced by the bilinear model. Scores are model confidences in [0, 1].

| Relation | Source | Target | Score | Status |
| --- | --- | --- | --- | --- |
| GENE_AFFECTS_DRUG | CYP2D6 | Codeine dosing adjustment | 0.842 | Supported [14] |
| VAR_ASSOC_DIS | HLA-B*15:02 | Carbamazepine-induced SJS/TEN | 0.817 | Supported [15] |
| DRUG_CAUSES_ADR | Simvastatin | Myopathy in SLCO1B1 c.521T>C carriers | 0.792 | Supported [16] |
| DRUG_IN_PATH | Metformin | AMPK-regulated autophagy pathway | 0.768 | Pending literature review |
| GENE_IN_PATH | CYP3A5 | Bile acid metabolism | 0.741 | Pending literature review |

### 3.4 Neighborhood Visualization

Figure 4 illustrates representative one-hop ego-graphs for top-scoring predictions, providing multi-relational evidence to support manual adjudication. Visualization dashboards enable interactive exploration with per-node provenance and timestamp metadata.

**Figure 4:**
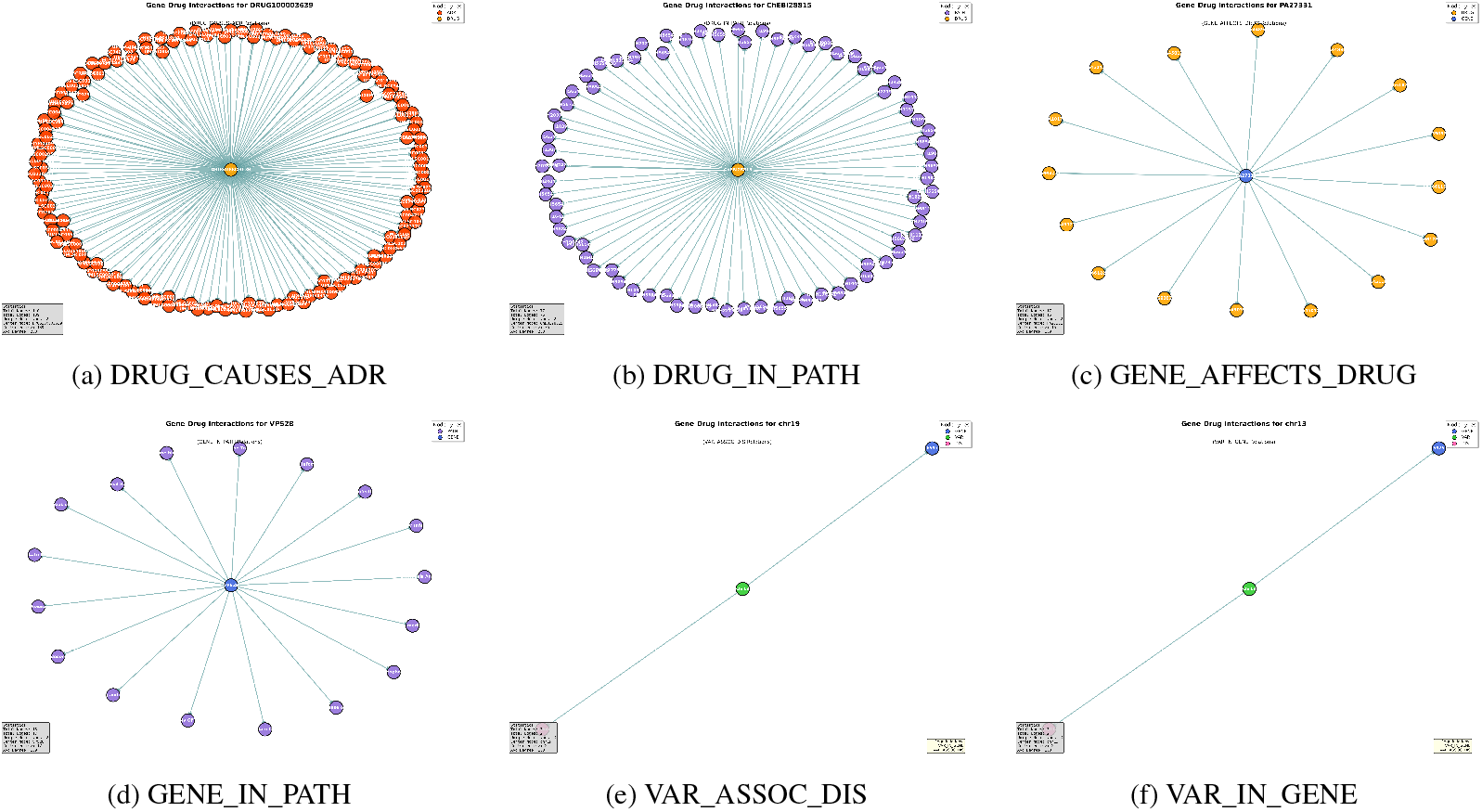
One-hop ego-graphs supporting selected predictions. Insets report per-graph summary statistics.

## 4 Discussion

The leakage-aware PGx-KG establishes a reproducible foundation for pharmacogenomic link prediction. Recovering guideline-backed associations demonstrates that the integration and auditing steps preserve clinically meaningful structure, while pending hypotheses provide a ranked queue for experimentalists. The bilinear baseline leaves headroom for more expressive relation-aware architectures, particularly on sparsely populated drug–pathway edges where performance remains modest.

### Implications for Precision Medicine

Integrating curated PGx resources supports downstream tasks such as adverse event mitigation, genotype-guided dosing, and target prioritization. Embedding-based ranking coupled with provenance-rich visualizations accelerates expert review by surfacing both molecular context and supporting literature.

### Future Work

Next steps include benchmarking CompGCN and inductive message-passing models, integrating real-world evidence streams (e.g., AEOLUS and EHR-derived signals), and expanding evaluation with causal discovery metrics. Public release will pair code, processed data, and documentation, enabling external validation and community-driven refinement.

## 5 Conclusion

PGx-KG combines rigorous data integration with leakage-free evaluation to advance pharmacogenomic hypothesis generation. By harmonizing multi-source evidence, enforcing temporal integrity, and establishing baseline performance, this work lays the groundwork for reproducible, large-scale discovery pipelines in precision medicine.

## Data Availability

All data produced in this study are available online. The processed pharmacogenomic knowledge graph (PGx-KG), including nodes, edges, relation-specific triples, benchmark splits, and metadata, has been deposited in Zenodo and is accessible at https://doi.org/10.5281/zenodo.17189995. All code and reproducibility materials are available at https://github.com/Faerque/pharmacogenomic-knowledge-graph. Raw source data are publicly available from PharmGKB, ClinVar, SIDER, and Reactome.

https://github.com/Faerque/pharmacogenomic-knowledge-graph

https://www.pharmgkb.org/

https://www.ncbi.nlm.nih.gov/clinvar/

http://sideeffects.embl.de/

https://reactome.org/

